# Improving inferences regarding individual patients using Emergency Medical Services response-based data in the National Emergency Medical Services Information System (NEMSIS)

**DOI:** 10.1101/2025.09.02.25334950

**Authors:** CN Morrison, BR Bushover, RP Crowe, CW Mills, AX Lo, AG Rundle

## Abstract

**Study Objective:** The National Emergency Medicine Services Information System (NEMSIS) public release dataset is an important tool for researching Emergency Medical Services (EMS) responses. However, multiple EMS units might attend to a single patient and the data are organized by EMS response, presenting challenges to inferences about patient-level events. We test whether data on time of the 911 call and patient characteristics can be used to screen for multiple EMS records that reflect a single patient encounter.

**Methods:** Using data on EMS responses to assaults in New York City in 2024 we identified EMS responses that had identical data for time of 911 call, patient age, sex, race/ethnicity, and longitude-latitude where the patient was encountered. EMS responses with identical data for all of these variables were assumed to have attended to a single patient. We then assessed the validity of using matches on 911 call time, patient age, sex, race/ethnicity (i.e., without latitude-longitude) to identify instances where separate EMS responses matched for these variables plus location.

**Results:** Of 32,202 EMS responses, 5,143 responses matched other responses for all variables, suggesting that there were 26,451 patients encounters. Matching on permutations of variables for time of 911 call, patient age, sex, race and ethnicity had 100% sensitivity and a high specificity (range 91.3% to 98.6%) for identifying responses that matched on all of these variables plus longitude-latitude.

**Conclusion:** Data available in the NEMSIS public release dataset can be used to screen for duplicate EMS responses improving inferences about patient level events.

## Introduction

### Background

The National Emergency Medicine Services Information System (NEMSIS) is the national database used to store Emergency Medical Services (EMS) data and provides a data format standard for how patient care information resulting from an emergency 911 call for assistance is collected, stored and distributed.^1,2^ NEMSIS serves as the primary data set used by researchers for pre-hospital health services research on EMS and is a valuable tool for measuring the burden and morbidity associated with injuries in the United States.^1,2^ The 2023 NEMSIS public release research dataset includes data on 54,190,579 EMS responses, from 14,369 EMS agencies serving 54 states and territories.^3^ The NEMSIS data are widely used in applications such as studying prehospital treatments, trends in the volume and types of EMS calls, and issues in medical transport; and have been proposed as a tool for national health surveillance for outdoor falls and domestic violence.^4-10^

### Importance

A common problem when examining EMS administrative data, including NEMSIS data, is identifying individual patients in the presence of double counting due to multiple ambulance responses for some patients.^1,11^ The data in the NEMSIS public release datasets are organized using the EMS activation or “response” as the unit of analysis. Data rows describe EMS responses and the database does not include a variable that identifies patients.^1^ Thus, if two EMS units respond to a single patient event, the patient-level event is represented twice in NEMSIS data as part of the data generated by each EMS unit.^1^ This data structure is consistent with the use of NEMSIS for health services planning for EMS. However, this data structure presents challenges when NEMSIS is used to infer information about patient-level events, such as measuring the morbidity and mortality burden of injuries, using NEMSIS data for health surveillance, investigations of prehospital interventions, and for understanding how medications are used during EMS responses.^7,12-15^ Fully identifiable patient-level data are available to EMS providers (e.g., name, home address) and matching algorithms that use these data systems to identify unique patients have demonstrated high sensitivity and specificity.^11^ However, the NEMSIS public release data set does not include identifiable patient-level data, so these methods are not available to users of this national resource.

Examples of issues that can arise in analyses of NEMSIS data include describing the burden of medical events by patient demographic characteristics or in understanding patient treatment patterns in the pre-hospital setting. ^10,13-17^ For instance, a recent analysis of NEMSIS data described EMS encounters for 309,442 pediatric behavioral health emergencies by patient sex and age.^16^ Odds ratios were presented for the use of restraints by patient age, evidence of alcohol use and place the patient was encountered.^16^ However, it is unclear how frequently multiple data rows appeared in NEMSIS for the same pediatric patient’s event as a result of multiple EMS units responding to the patient. An example of ambiguity in analyses of pre-hospital treatment can be seen in the literature on the administration of naloxone to opioid overdose patients. ^10,13-15,17^ If an opioid overdose patient is attended to by two EMS units, and one unit administers naloxone, that single administration can be referenced in the data that each of the two units report to NEMSIS.^1^ That is, a report of naloxone administration will appear in NEMSIS for two EMS responses, but only one patient received naloxone. This issue likely effects estimates of the percent of EMS responses for opioid overdoses that involve naloxone administration and analyses of the frequency of which naloxone is administered multiple times to a single opioid overdose Patient_10,13-15,17_

### Goals of This Investigation

To improve the field’s ability to understand patient-level events and characteristics in the NEMSIS data, we propose, and validate, a method to use data elements in the public release NEMSIS datasets to identify occurrences of multiple EMS units attending to the same patient event. We hypothesized that when data on the time of 911 call, patient age, sex, race and ethnicity match across different entries in NEMSIS, the data entries represented multiple EMS units attending to the same patient.

## Methods

### Study Design and Setting

We sought to identify records in EMS administrative data where the day and time of the 911 call, patient age, sex, race and ethnicity and location where the patient was encountered were identical across multiple records in the data set. Records that matched on these variables were presumed to represent instances in which multiple EMS units responded to the same patient. We then tested the validity of using just data on time of the 911 call, patient age, sex, race and ethnicity – data fields within the public release NEMSIS datasets – to identify the records that matched on these variables plus location.

As the NEMSIS national research dataset does not contain any identifying information for patients or the location EMS responded to, we leveraged EMS administrative records from the New York State Department of Health (NYSDOH) from 2024, that do include data on the location where patients were encountered. This analysis leveraged a dataset used for another project on assault patients treated by EMS New York City. Since there is a large volume of EMS responses to assaults in NYC, we expected that these data would provide a suitably challenging context in which to test matching based strategies to identify multiple EMS units that responded to the same patient.

All records for EMS ground (non-helicopter) responses in New York are collected and stored using the NEMSIS data standard and are submitted to the NEMSIS repository. The NYSDOH provided EMS response-level data for all events with ICD-10 codes X93 (assault by handgun discharge), X94 (assault by rifle, shotgun and larger firearm discharge), X95 (assault by other and unspecified firearm and gun discharge), X99 (assault by sharp object), and Y04 (assault by bodily force). NEMSIS standards variables provided by NYSDOH were eInjury.01 (cause of injury code), eInjury.02 (mechanism of injury), ePatient.13 (gender), ePatient.14 (race/ethnicity), ePatient.15 (age), ePatient.16 (age units), eScene.11 (scene GPS coordinates), eTimes.01 (date and time of the call to a public safety answering service (PSAP)).

This study was reviewed by the Columbia University Irving Medical Center Institutional Review Board and classified as “Not Human Subjects Research Under 45 CFR 46”

### Measures

We created a “gold standard” definition for multiple EMS units responding to the same patient event as: EMS records that had the same PSAP call day and hour; the same longitude and latitude at three decimal places for the location where the patient was encountered (approximately 100 meters in New York City); and the same reported patient sex, age, race and ethnicity.

We then tested whether identifying records that matched for various combinations of the variables available in the national public use NEMSIS research dataset could be used to replicate the “gold standard” definition of multiple EMS units responding to the same patient event. The combinations of variables were:

Match Algorithm 1: same PSAP call day and hour, patient sex, age, race and ethnicity

Match Algorithm 2: same PSAP call day and hour, patient sex, and age

Match Algorithm 3: same PSAP call day and hour, and patient sex

Match Algorithm 4: same PSAP call day and hour, and patient age

### Analysis

The number of putative unique patient events under each Match Algorithm was compared to the number of putative unique patient events identified by the “gold standard” classifier. In addition, each EMS response record was classified as being for a unique patient or being for a non-unique patient under each Match Algorithm. The sensitivity, specificity, positive predictive value and negative predictive value was calculated for each Match Algorithm predicting the non-unique compared to the unique classifications created by the “gold standard” classifier.

## Results

The EMS administrative record data from the NYSDOH included 34,806 EMS responses for assaults. We excluded 2,604 (7%) responses that were missing information for the variables of interest: sex (109 responses), race/ethnicity (2 responses), age (22 responses), event date and hour (29 responses), and latitude-longitude (2,534 responses). The final analytic sample included 32,202 responses that comprised all EMS responses for assaults in New York City in 2024 with complete data.

In total the “gold standard” matching criteria indicated that there were 26,451 unique patient events: 21,308 of whom had 1 EMS response, 4,540 had 2 EMS responses, 600 had 3 EMS responses, 2 had 4 EMS responses, 0 had 5 EMS responses, and 1 had 6 EMS responses. Thus, there were a total of 5,143 patient events (19.4% of unique patient events) where it is presumed that more than one EMS unit responded to the patient. While the “gold standard” algorithm utilized matching on the first three decimal places of the longitude and latitude data, inspection of the data showed that all of the matched responses had the same longitude and latitude to six decimal places. The number of unique patients identified using each Match Algorithm was very close (within 4.6%) to the number of unique patients identified using the “gold standard” classifier (table 1), with Match Algorithm 1 having the smallest percent difference. Match Algorithm 1 identified 26,255 unique patient events, which was 196 (0.7%) fewer compared to the “gold standard” algorithm.

**Table 1.**
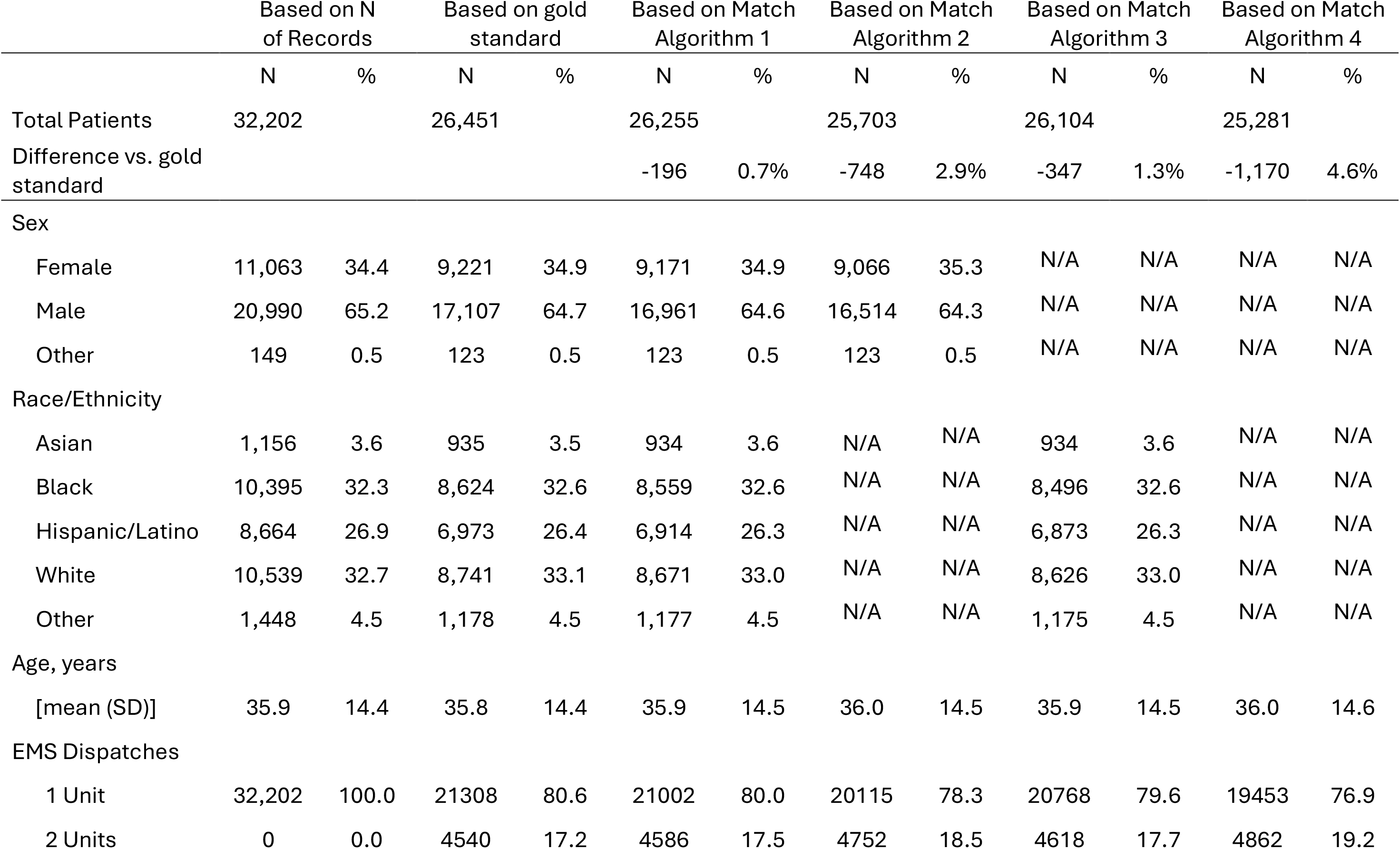
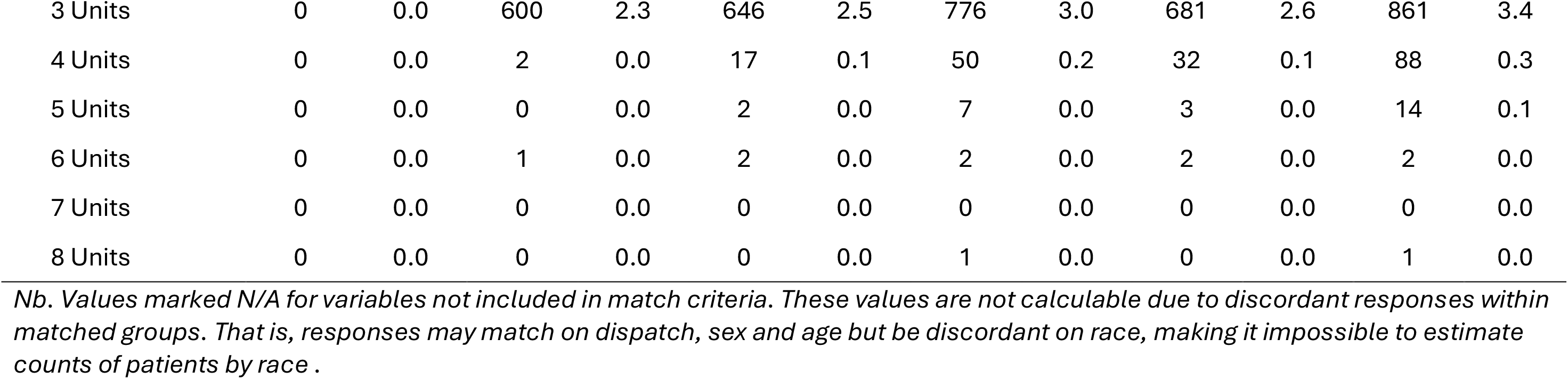
Descriptive statistics according to matching algorithm for identifying unique patient encounters

|  | Based on N<br>of Records |  | Based on gold<br>standard |  | Based on Match<br>Algorithm 1 |  | Based on Match<br>Algorithm 2 |  | Based on Match<br>Algorithm 3 |  | Based on Match<br>Algorithm 4 |  |
| --- | --- | --- | --- | --- | --- | --- | --- | --- | --- | --- | --- | --- |
|  | N | % | N | % | N | % | N | % | N | % | N | % |
| Total Patients | 32,202 |  | 26,451 |  | 26,255 |  | 25,703 |  | 26,104 |  | 25,281 |  |
| Difference vs. gold<br>standard |  |  |  |  | -196 | 0.7% | -748 | 2.9% | -347 | 1.3% | -1,170 | 4.6% |
| Sex |  |  |  |  |  |  |  |  |  |  |  |  |
| Female | 11,063 | 34.4 | 9,221 | 34.9 | 9,171 | 34.9 | 9,066 | 35.3 | N/A | N/A | N/A | N/A |
| Male | 20,990 | 65.2 | 17,107 | 64.7 | 16,961 | 64.6 | 16,514 | 64.3 | N/A | N/A | N/A | N/A |
| Other | 149 | 0.5 | 123 | 0.5 | 123 | 0.5 | 123 | 0.5 | N/A | N/A | N/A | N/A |
| Race/Ethnicity |  |  |  |  |  |  |  |  |  |  |  |  |
| Asian | 1,156 | 3.6 | 935 | 3.5 | 934 | 3.6 | N/A | N/A | 934 | 3.6 | N/A | N/A |
| Black | 10,395 | 32.3 | 8,624 | 32.6 | 8,559 | 32.6 | N/A | N/A | 8,496 | 32.6 | N/A | N/A |
| Hispanic/Latino | 8,664 | 26.9 | 6,973 | 26.4 | 6,914 | 26.3 | N/A | N/A | 6,873 | 26.3 | N/A | N/A |
| White | 10,539 | 32.7 | 8,741 | 33.1 | 8,671 | 33.0 | N/A | N/A | 8,626 | 33.0 | N/A | N/A |
| Other | 1,448 | 4.5 | 1,178 | 4.5 | 1,177 | 4.5 | N/A | N/A | 1,175 | 4.5 | N/A | N/A |
| Age, years |  |  |  |  |  |  |  |  |  |  |  |  |
| [mean (SD)] | 35.9 | 14.4 | 35.8 | 14.4 | 35.9 | 14.5 | 36.0 | 14.5 | 35.9 | 14.5 | 36.0 | 14.6 |
| EMS Dispatches |  |  |  |  |  |  |  |  |  |  |  |  |
| 1 Unit | 32,202 | 100.0 | 21308 | 80.6 | 21002 | 80.0 | 20115 | 78.3 | 20768 | 79.6 | 19453 | 76.9 |
| 2 Units | 0 | 0.0 | 4540 | 17.2 | 4586 | 17.5 | 4752 | 18.5 | 4618 | 17.7 | 4862 | 19.2 |
| 3 Units | 0 | 0.0 | 600 | 2.3 | 646 | 2.5 | 776 | 3.0 | 681 | 2.6 | 861 | 3.4 |
| 4 Units | 0 | 0.0 | 2 | 0.0 | 17 | 0.1 | 50 | 0.2 | 32 | 0.1 | 88 | 0.3 |
| 5 Units | 0 | 0.0 | 0 | 0.0 | 2 | 0.0 | 7 | 0.0 | 3 | 0.0 | 14 | 0.1 |
| 6 Units | 0 | 0.0 | 1 | 0.0 | 2 | 0.0 | 2 | 0.0 | 2 | 0.0 | 2 | 0.0 |
| 7 Units | 0 | 0.0 | 0 | 0.0 | 0 | 0.0 | 0 | 0.0 | 0 | 0.0 | 0 | 0.0 |
| 8 Units | 0 | 0.0 | 0 | 0.0 | 0 | 0.0 | 1 | 0.0 | 0 | 0.0 | 1 | 0.0 |

*Nb. Values marked N/A for variables not included in match criteria. These values are not calculable due to discordant responses within matched groups. That is, responses may match on dispatch, sex and age but be discordant on race, making it impossible to estimate counts of patients by race .*

When each EMS response was classified using the “gold standard” as being for a unique or non-unique patient, 21,308 responses were for unique patients and 10,894 responses (34%, see Table 2) were for non-unique patients (5,143 unique patients). Classifications created using each of the Match Algorithms were highly predictive of the “gold standard” classification. Compared to the “gold standard” classifier, all four of the Match Algorithms had 100% sensitivity for identifying duplicate records and had high specificity, with Match Algorithm 1 having the highest specificity (98.6%) and highest negative predictive value (97.3%).

**Table 2.** Diagnostic performance of Matching Algorithms 1-4 for identifying duplicate EMS responses compared to those found using the “gold standard” matching algorithm.

| Match Test | EMS Responses to Unique Patients | EMS Responses to Non-Unique Patients | Sensitivity | Specificity | Positive Predictive Value | Negative Predictive Value |
| --- | --- | --- | --- | --- | --- | --- |
| Gold Standard | 21,308 | 10,894 |  |  |  |  |
| Match Algorithm 1 | 21,002 | 11,200 | 100 | 98.6 | 100 | 97.3 |
| Match Algorithm 2 | 20,115 | 12,087 | 100 | 94.4 | 100 | 90.1 |
| Match Algorithm 3 | 20,768 | 11,434 | 100 | 97.5 | 100 | 95.3 |
| Match Algorithm 4 | 19,453 | 12,749 | 100 | 91.3 | 100 | 85.5 |

## Discussion

Matching data from EMS activations on PSAP call day and hour, location where the patient was encountered and patient sex, age and race and ethnicity suggested that there were a substantial number of instances where multiple EMS units responded to a single assault patient. While data on longitude and latitude of the location of the patient encounter are not available in the public release NEMSIS data, the analyses presented here show that matching on variables that are available in the public release data sets identifies EMS responses that can be presumed to have attended to the same patient. We argue that applying these matching procedures in an analysis of the public release NEMSIS data allows for improved estimation of patient-level characteristics and pre-hospital treatment patterns from EMS response-level data. At the least these matching procedures to identify putative non-unique patients allow for credible bounds to be placed upon inferences or statements regarding patient-level events or characteristics.

The analyses presented here have some limitations that should be considered. The analyses focused on a frequently occurring 911 call type in a single densely populated city. For nation-wide analyses there may be sufficient numbers of patients that matching on PSAP call day and hour, patient sex, age, race and ethnicity does not identify unique patients. Implementations of this matching based method in nation-wide analyses should also match on Census region and perhaps minute of the PSAP call. In addition, we are conceptualizing this matching approach as a screening test, shown here to have a high sensitivity and specificity, for identifying multiple EMS responses to unique patient events. However, the influence of the prevalence of the outcome (e.g. “gold standard” classification as unique or non-unique patient) needs to be considered when positive and negative predictive values are used to judge the quality of a screening test (e.g. Match Algorithm 1).^18^ For a given sensitivity and specificity, the negative predictive value of a screening test is higher for a low prevalence outcome and is lower for a high prevalence outcome.^18^ There are dispatch call types (e.g. out of hospital cardiac event) where it is common for more than one EMS unit to attend to a patient and dispatch call types where it is less common for more than one EMS unit to attend to a patient (e.g. opioid overdoses). Thus, call type should be considered when interpreting the results of this approach to identifying EMS responses that attended to the same patient. Again, we argue that the method presented here should be conceptualized as a way to place realistic bounds on estimates of numbers and rates of patient-level events or characteristics inferred from EMS response-level data in NEMSIS.

Our use of granular EMS data from New York City included data not available through the publicly available NEMSIS dataset, which allowed for our validation analyses. It is possible that data from New York City may not be generalizable to analyses of NEMSIS data from other locations. However, we would not expect any systematic biases involving the variables and data selected for use in our matching algorithms across locations, and therefore anticipate that our findings would be applicable to NEMSIS data across the U.S.

In conclusion, matching on PSAP day and hour and patient characteristics, along with region, appears to be a viable way to identify in the public release NEMSIS data occurrences of multiple EMS units attending to the same patient. Implementation of this approach will strength inferences about patient-level characteristics, events, counts and rates from analyses of the NEMSIS data.

## Data Availability

Data are available from the New York State Department of Health

## Acknowledgments

Drs. Morrison and Rundle were supported in part by Emergent BioSolutions through a contract with Columbia University. Drs. Morrison and Rundle were also provided support by Columbia Center for Injury Science and Prevention, Centers for Disease Control and Prevention (CDC) grant no. R49CE003094. Dr. Crowe is a current employee of ESO Solutions. Dr. Mills is a current employee of Emergent BioSolutions and holds equity in the company.

